# Tracking Progress: Trends in Surgical Indicator Reporting in the World Development Indicators

**DOI:** 10.1101/2024.12.24.24316011

**Authors:** Carolina Torres Perez-Iglesias, Kennedy Jensen, Vanitha Raguveer, Gabriella Y. Hyman, Emi Suzuki, John G. Meara, Nakul P. Raykar

**Affiliations:** Program in Global Surgery and Social Change, Blavatnik Institute Department of Global Health and Social Medicine, Harvard Medical School; University of Utah Department of General Surgery; Department of Surgery, University of the Witwatersrand; Development Data Group, World Bank; Boston Children’s Hospital

## Abstract

**Background:** The Lancet Commission on Global Surgery (LCoGS) emphasized the need for six core indicators to track progress toward safe, affordable, and timely surgical care. In 2016, four of these indicators were integrated into the World Development Indicators (WDI) to facilitate global tracking of surgical system performance. However, the impact of these indicators relies on the accuracy and frequency of the reported data. We aimed to analyze trends in the reporting of the WDI surgical indicators and evaluate trends in the context of countries undertaking National Surgical Plans (NSPs).

**Methods:** Data were extracted from the World Bank WDI database and analyzed by WHO region from 2015-2020. Countries were categorized based on their engagement with NSPs. Trends in reporting for the four surgical indicators—surgical case volume (SV), surgical workforce density (SW), catastrophic health expenditure (CHE), and impoverishing expenditure (IE)—were assessed. Correlations between NSP implementation and reporting completeness were explored.

**Results:** From 2015 to 2020, reporting rates for the modeled indicators CHE and IE remained stable, with an average annual reporting rate of 62% for both. In contrast, the reporting of non-modeled indicators (SV, SW) declined over the study period. SV reporting dropped from 12% in 2015 to 0% in 2020, and SW reporting decreased from 18% to 0%. Regional disparities were significant: the European region had the highest reporting rates for non-modeled indicators, with 13.3% for SW and 3.7% for SV, while the African region reported only 5.4% for SW and 1.8% for SV. No significant correlation was found between NSP implementation and reporting rates (R² = 0.00014, p = 0.986).

**Conclusions:** Despite integrating surgical indicators into the WDI, reporting rates remain low, particularly for non-modeled indicators. Strengthening data collection and reporting systems, particularly in low-resource settings, is essential for improving surgical care monitoring and addressing global disparities.

## INTRODUCTION

The Lancet Commission on Global Surgery (LCoGS), published in 2015, demonstrated the importance of six core indicators essential for measuring universal access to safe, timely, affordable, and equitable surgical care. (1) When considered together, these indicators provide a framework for understanding surgical health system functioning and addressing disparities in surgical care provision. These indicators can be used to monitor and evaluate global surgical initiatives and identify areas for improvement. (2) However, the effectiveness of these indicators in providing an accountable and comprehensive view of surgical care depends heavily on the accuracy and consistency of the data reported.

Following a collaborative effort to gather indicator data globally, four of the six consensus-based LCoGS indicators were integrated into the World Bank’s World Development Indicators (WDI) in 2016, including 1) *Surgical Volume (Number of surgical procedures per 100,000 population)*, 2) *Surgical Specialist Workforce Density* (Specialist surgical workforce per 100,000 population), 3) Protection Against Impoverishing Expenditure (*Percentage of Population at Risk of Impoverishing Expenditure for Surgical Care)*, and 4) Protection Against Catastrophic Expenditure (*Percentage of Population at Risk of Catastrophic Expenditure for Surgical Care*). (3) The original LCoGS report intended for these metrics to be tracked and reported by all countries and global health organizations. Since its publication, some countries have collected and reported local indicator data, however, these have not been universally integrated into national health data systems. (4–6) To aid with the integration of surgical indicators in national health systems, the LCoGS proposed the development of national surgical plans, also known as National Surgical Obstetric and Anaesthesia Plans (NSOAPs). These plans were designed to serve as strategic frameworks to integrate surgical care into national health systems by identifying and addressing gaps in surgical care within a country.

Since their inception and integration into the WDI database, the six LCoGS indicators have undergone iterative validation processes and have been consolidated into five indicators. (7,8) As we approach ten years after the publication of the surgical indicators, there is a need to evaluate progress both in terms of reaching the targets set out in LCoGS and in reporting the surgical indicators used to inform health policy and practice. We aimed to assess the trends in reporting of surgical indicators in the World Bank’s WDI and to evaluate these trends in the context of countries undertaking national surgical plans.

## METHODS

### Context

The LCoGS proposed six indicators divided into three groups. Box 1 defines these six indicators along with each of these originally suggested data sources, targets for 2030, and method of reporting in WDI currently used. Responsible entities for the collection of these indicators included local agencies such as the Ministry of Health, healthcare facilities, and household surveys.

#### Box 1

Original six core indicators defined by the LCoGS including proposed data sources and targets by 2030 (1)

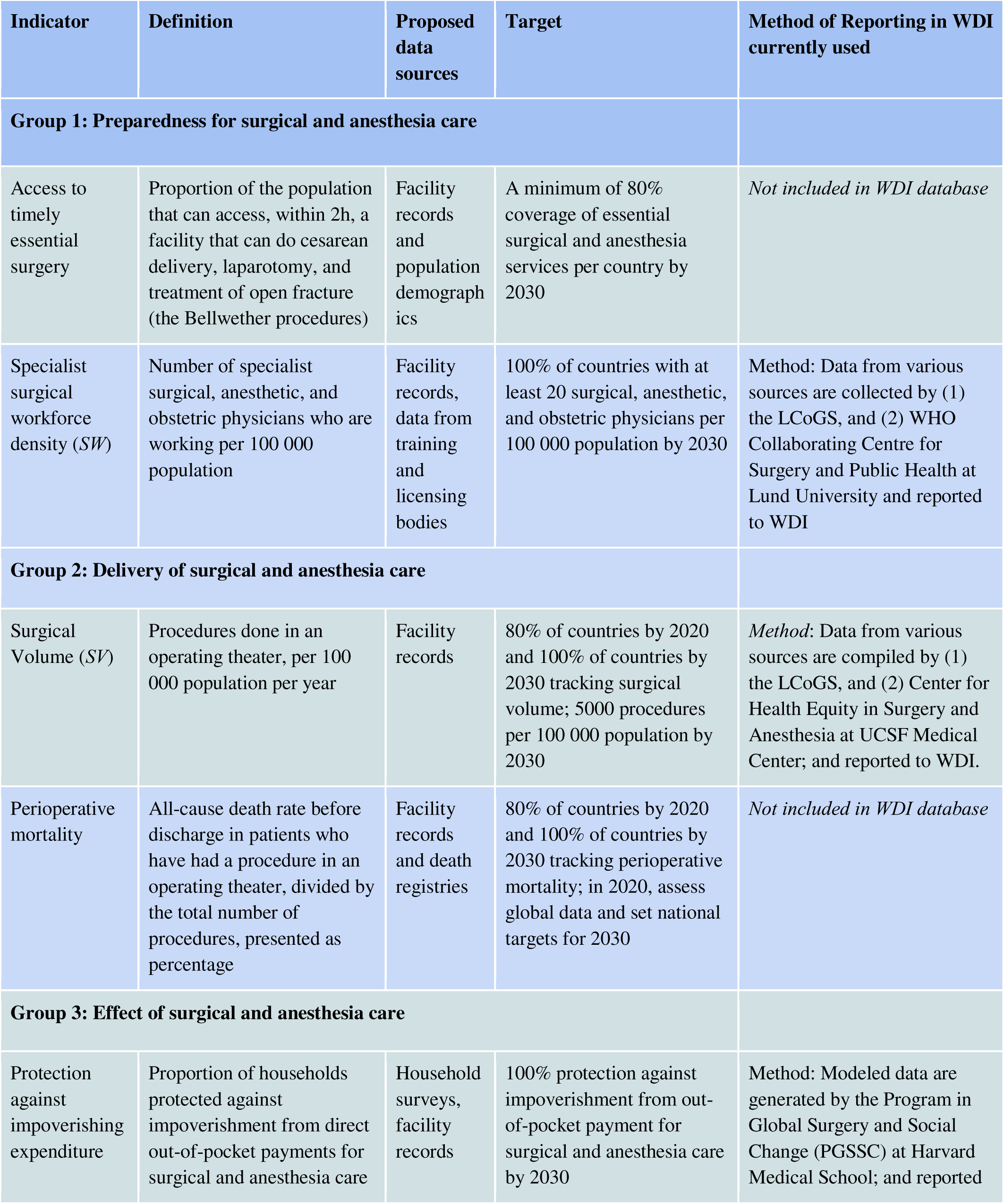
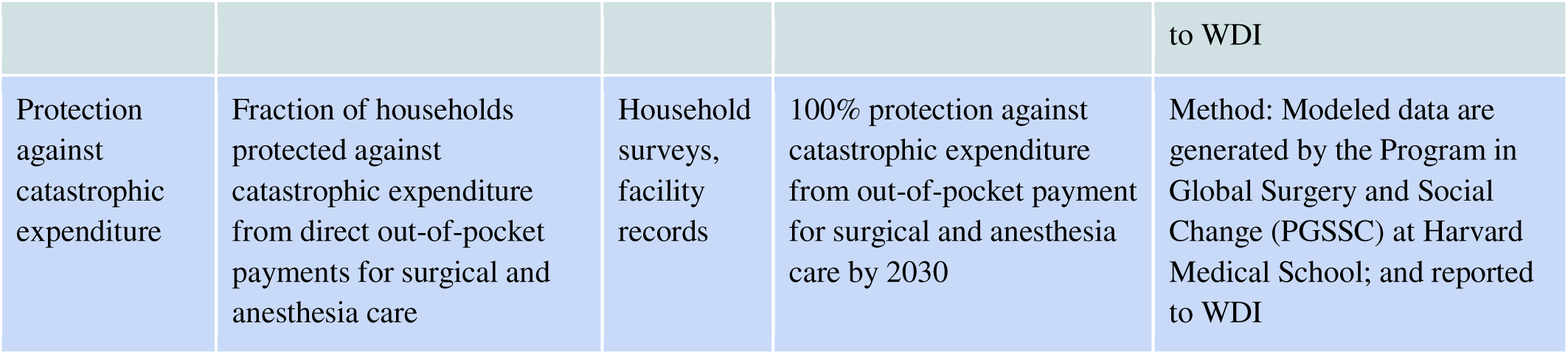

**Box 1: Original six core indicators defined by the LCoGS including proposed data sources and targets by 2030 (1)**
| Indicator | Definition | Proposed data sources | Target | Method of Reporting in WDI currently used |
| --- | --- | --- | --- | --- |
| **Group 1: Preparedness for surgical and anesthesia care** |  |  |  |  |
| Access to timely essential surgery | Proportion of the population that can access, within 2h, a facility that can do cesarean delivery, laparotomy, and treatment of open fracture (the Bellwether procedures) | Facility records and population demographics | A minimum of 80% coverage of essential surgical and anesthesia services per country by 2030 | *Not included in WDI database* |
| Specialist surgical workforce density (SW) | Number of specialist surgical, anesthetic, and obstetric physicians who are working per 100 000 population | Facility records, data from training and licensing bodies | 100% of countries with at least 20 surgical, anesthetic, and obstetric physicians per 100 000 population by 2030 | Method: Data from various sources are collected by (1) the LCoGS, and (2) WHO Collaborating Centre for Surgery and Public Health at Lund University and reported to WDI |
| **Group 2: Delivery of surgical and anesthesia care** |  |  |  |  |
| Surgical Volume (SV) | Procedures done in an operating theater, per 100 000 population per year | Facility records | 80% of countries by 2020 and 100% of countries by 2030 tracking surgical volume; 5000 procedures per 100 000 population by 2030 | *Method:* Data from various sources are compiled by (1) the LCoGS, and (2) Center for Health Equity in Surgery and Anesthesia at UCSF Medical Center; and reported to WDI. |
| Perioperative mortality | All-cause death rate before discharge in patients who have had a procedure in an operating theater, divided by the total number of procedures, presented as percentage | Facility records and death registries | 80% of countries by 2020 and 100% of countries by 2030 tracking perioperative mortality; in 2020, assess global data and set national targets for 2030 | *Not included in WDI database* |
| **Group 3: Effect of surgical and anesthesia care** |  |  |  |  |
| Protection against impoverishing expenditure | Proportion of households protected against impoverishment from direct out-of-pocket payments for surgical and anesthesia care | Household surveys, facility records | 100% protection against impoverishment from out-of-pocket payment for surgical and anesthesia care by 2030 | Method: Modeled data are generated by the Program in Global Surgery and Social Change (PGSSC) at Harvard Medical School; and reported |

|  |  |  |  | to WDI |
| --- | --- | --- | --- | --- |
| Protection against catastrophic expenditure | Fraction of households protected against catastrophic expenditure from direct out-of-pocket payments for surgical and anesthesia care | Household surveys, facility records | 100% protection against catastrophic expenditure from out-of-pocket payment for surgical and anesthesia care by 2030 | Method: Modeled data are generated by the Program in Global Surgery and Social Change (PGSSC) at Harvard Medical School; and reported to WDI |

### Study design

The WDI is a comprehensive dataset provided by the World Bank, which includes a range of economic, social, environmental, and demographic indicators collected globally and used to assess and compare global development trends and progress over time.

The WDI includes four of the original six LCoGS indicators:

- Number of surgical procedures (per 100,000 population) (SV)
- Specialist surgical workforce (per 100,000 population) (SW)
- Risk of catastrophic expenditure for surgical care (% of people at risk) (CE)
- Risk of impoverishing expenditure for surgical care (% of people at risk) (IE).

Data from the four surgical indicators was accessed through the World Bank DataBank in May 2024. This open-access online platform provides access to multiple datasets collected by the World Bank Group, including the WDI. Data was collected from 2015 to 2020, including 194 United Nations Member States. Countries were categorized by WHO region (Africa, Eastern Mediterranean, Europe, Americas, South-East Asia, Western Pacific). (9) Non-member states and territories were excluded. Data after 2020 was excluded to mitigate the effect the COVID-19 pandemic may have had as a potential confounder in data collection and reporting. Additionally, data beyond this range was not available at the time of the analysis. Data was obtained from the open-access World Development Indicators DataBank. (10)

Data on the development and implementation of national surgical plans (NSPs), also known as national surgical, obstetric, and anesthesia plans (NSOAPs), was gathered through an extensive internet-based search, which included academic literature (peer-reviewed publication and gray literature), publicly available documentation, published reports and supplemented data from global health databases and registries.

### Statistical analysis

Descriptive statistics were used to analyze the trends in reporting rates for each indicator. Completeness of data and response rates were assessed by region. A trend analysis across regions was performed to identify patterns, outliers, and changes over the study period. Additionally, we examined correlations with the implementation of NSPs. All data was open-source and freely available to the general public. No IRB approval was requested for this study.

## RESULTS

Throughout the study period, data reporting for surgical indicators varied considerably across WHO regions, with high rates of missing values noted, suggesting incomplete data availability, collection or inconsistencies in reporting over time. Financial risk indicators were the most consistently reported across all regions and years. In contrast, SV and SW displayed substantial gaps and declining reporting over the years. Table 1 presents the reporting rates of each indicator across WHO region from 2015 to 2020, highlighting the presence of incomplete data for several indicators and years. (--indicates *no data reported*).

**Table 1:**
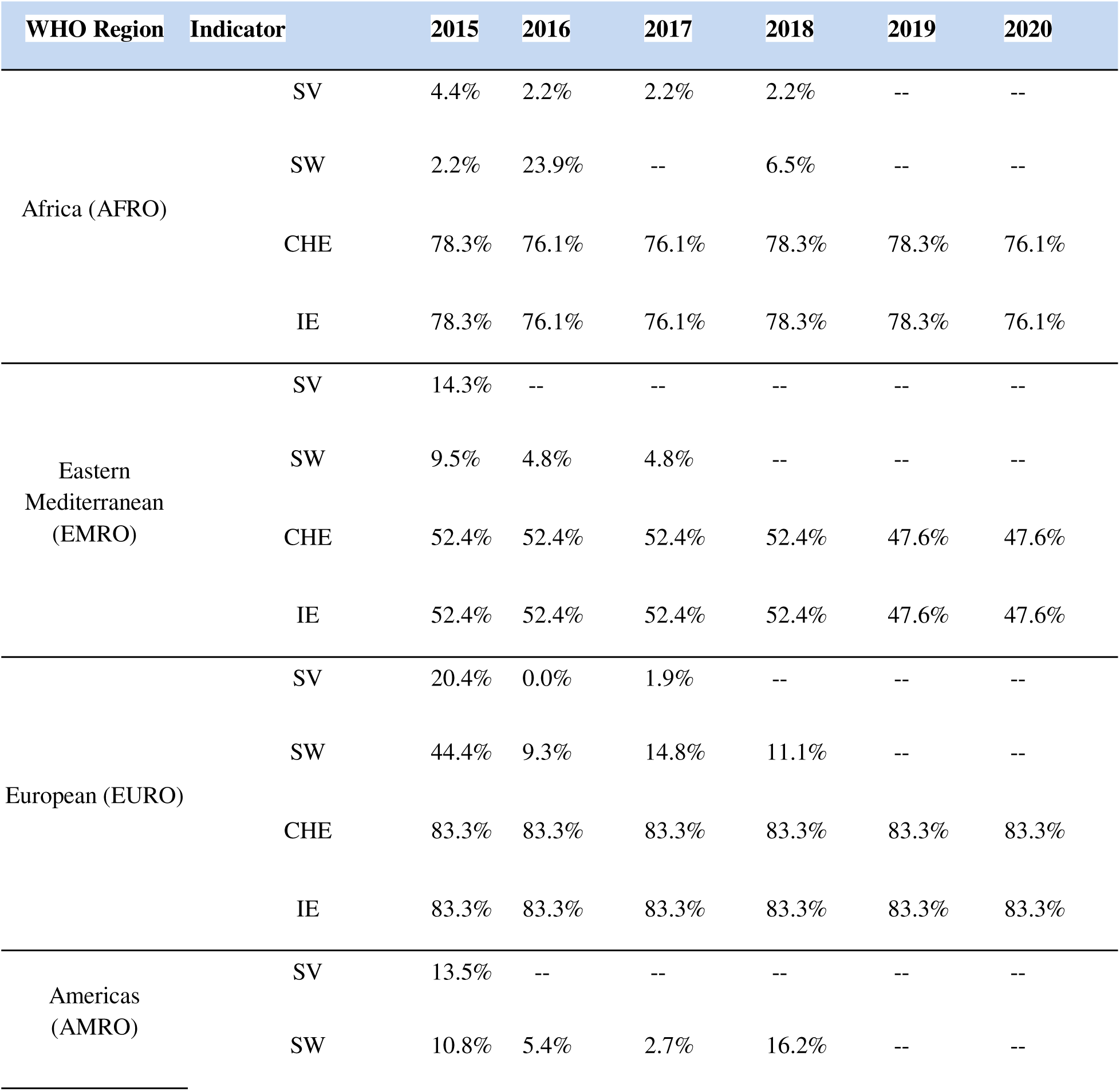
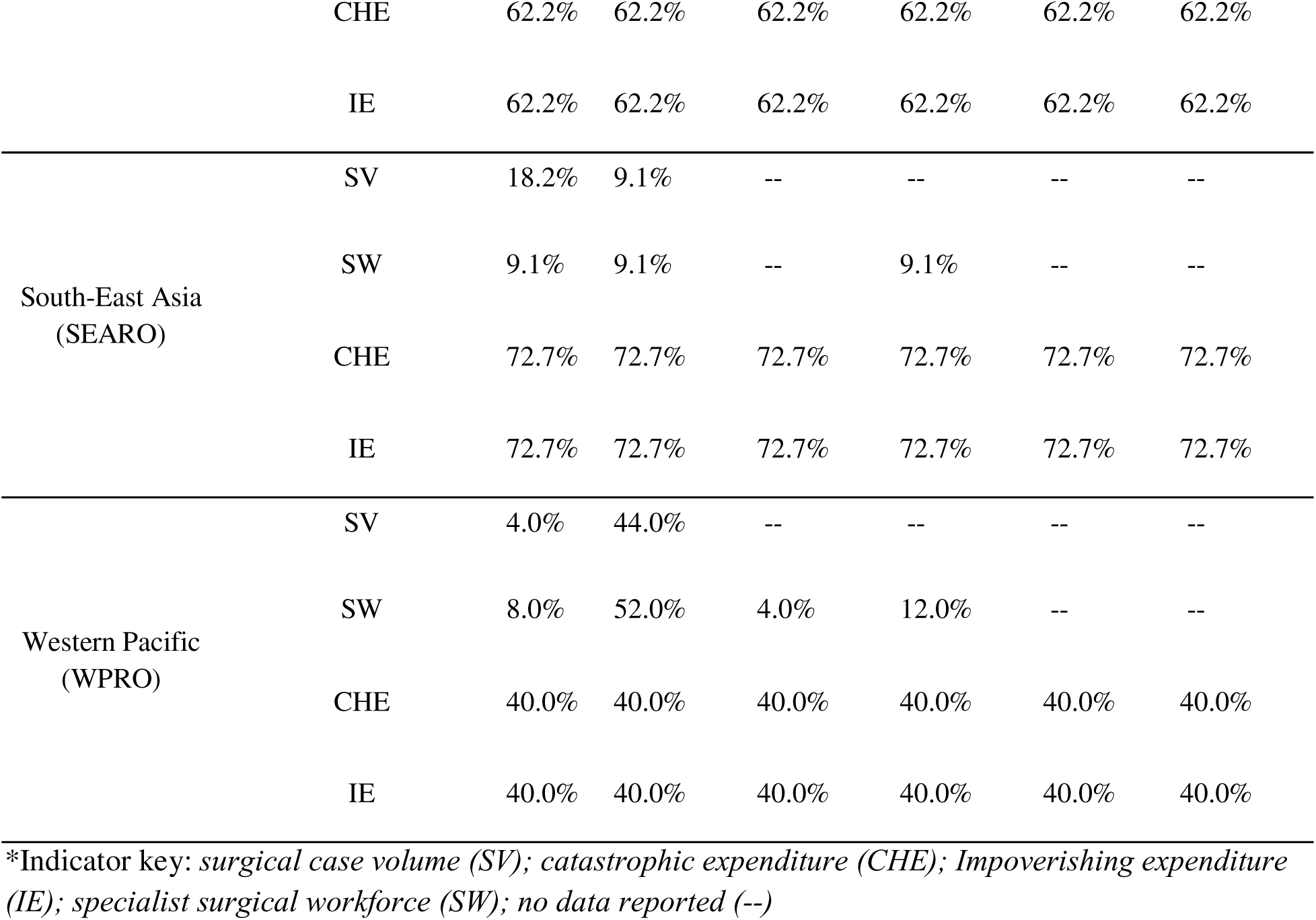
Surgical indicator reporting rates (%) across WHO regions, from 2015 to 2020.

### Indicator Trends

Within the study period, one or more surgical indicators were reported for 157 (81%) countries in the WDI at some point. The financial indicators, CHE and IE, both of which are modeled indicators and do not require reporting on the part of the country, were reported at the highest rates (median 100%; IQR 100%) and remained consistent overall, with between 61% and 62% reporting annually. Reporting for non-modeled indicators, SV and SW, decreased from 12% and 18%, respectively, in 2015 to 0% in 2020 (SV MED=0, IQR=0; SW MED=0, IQR=16.7%). Regional differences in reporting rates were noted within the dataset. The European and Western Pacific regions demonstrated the highest reporting rates for the non-modeled indicators: 13.3% and 12.7% for SW, 3.7% and 8.0% for SV, respectively. In contrast, the African region exhibited the second lowest reporting rates globally, averaging 1.8% for SV and 5.4% for SW over the study period (Figure 1, Figure 2).

**Figure 1:**
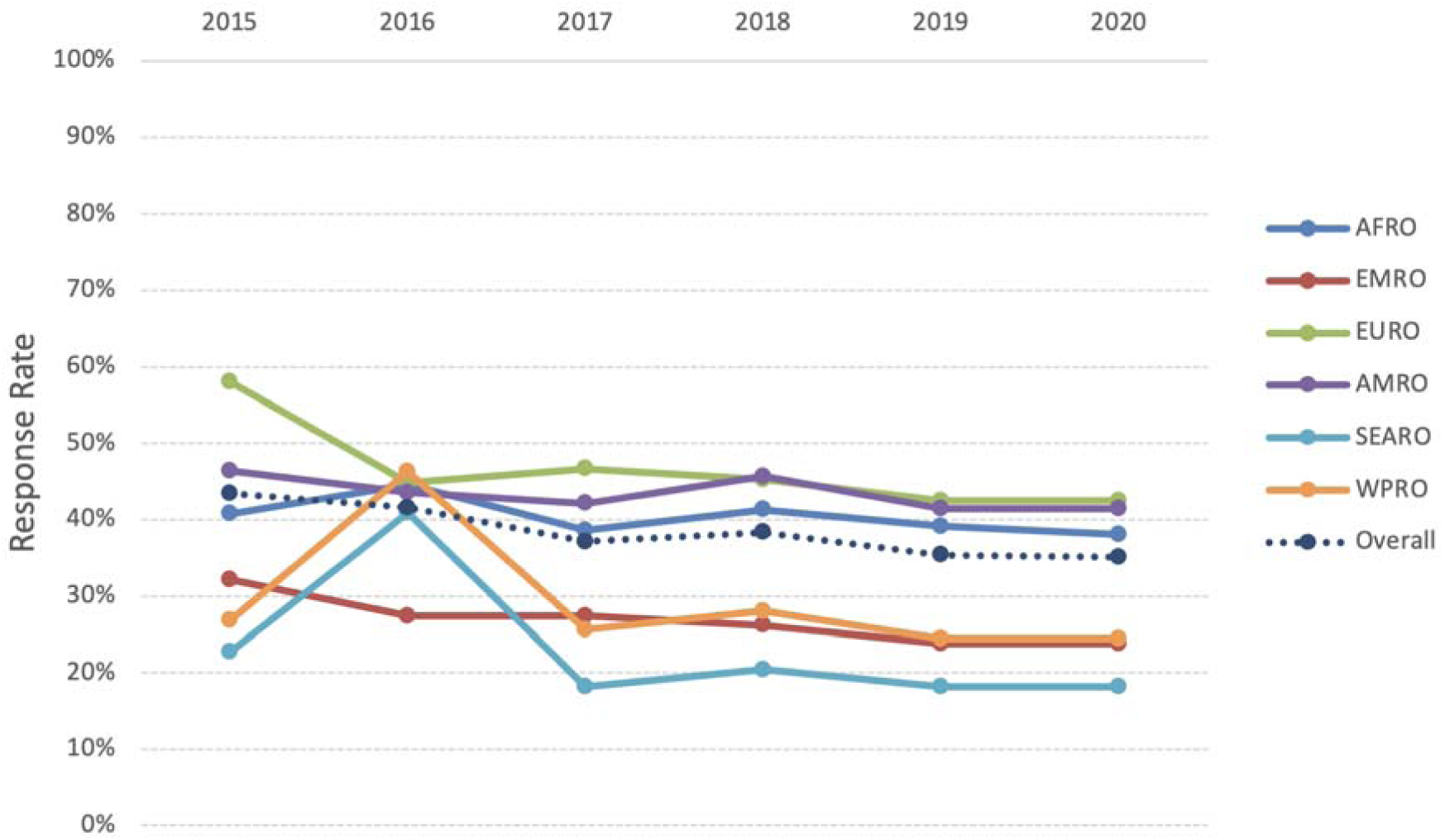
Trends in Average Reporting Rates for All Surgical Indicators by WHO Region.

**Figure 2:**
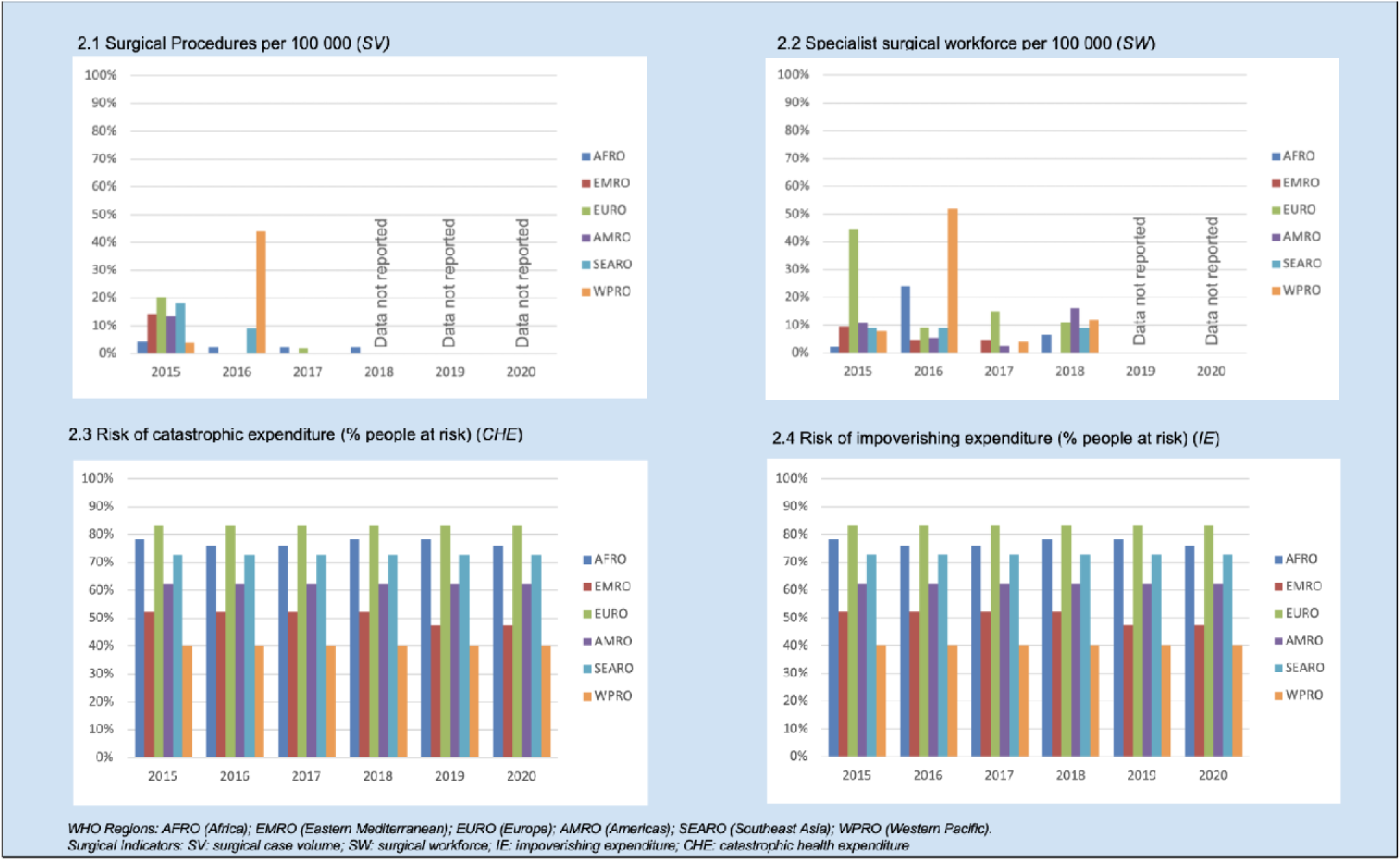
Trends in surgical indicator reporting by WHO Region from 2015-2020.

### NSP Trends

We used NSP development as a proxy for country-level commitment to surgical system strengthening and data collection. Many countries around the world have undertaken NSP development. Most of these countries are in the WHO AFRO Region, followed by WPRO. However, undertaking NSP development did not correlate with the reporting of surgical indicators for the WDI (Figure 3). Multiple linear regression was used to test if NSP implementation or development predicted indicator reporting rates from 2015-2020. The overall regression was not statistically significant (R^2^ = 0.00014, F(2,191) = 0.0137, p = 0.986). Neither NSP implementation (β = 0.005, p = 0.95) nor NSP development of any stage (β = 0.005, p = 0.93) significantly predicted reporting rates. In the Western Pacific Region, ten countries have undertaken NSPs, while 13 countries reported non-modeled indicators during the study period, 8 (62%) of which were NSP-engaged countries. In the African region, 16 countries are engaged in the NSP process, with 15 countries reporting non-modeled indicators, 9 (60%) of which are NSP engaged. Seven of the nine countries in the NSP implementation phase are located in the African Region, with indicator reporting peaking at 64% in 2016 and subsequently declining to 43% for 2017-2020 for these implementing countries.

**Figure 3:**
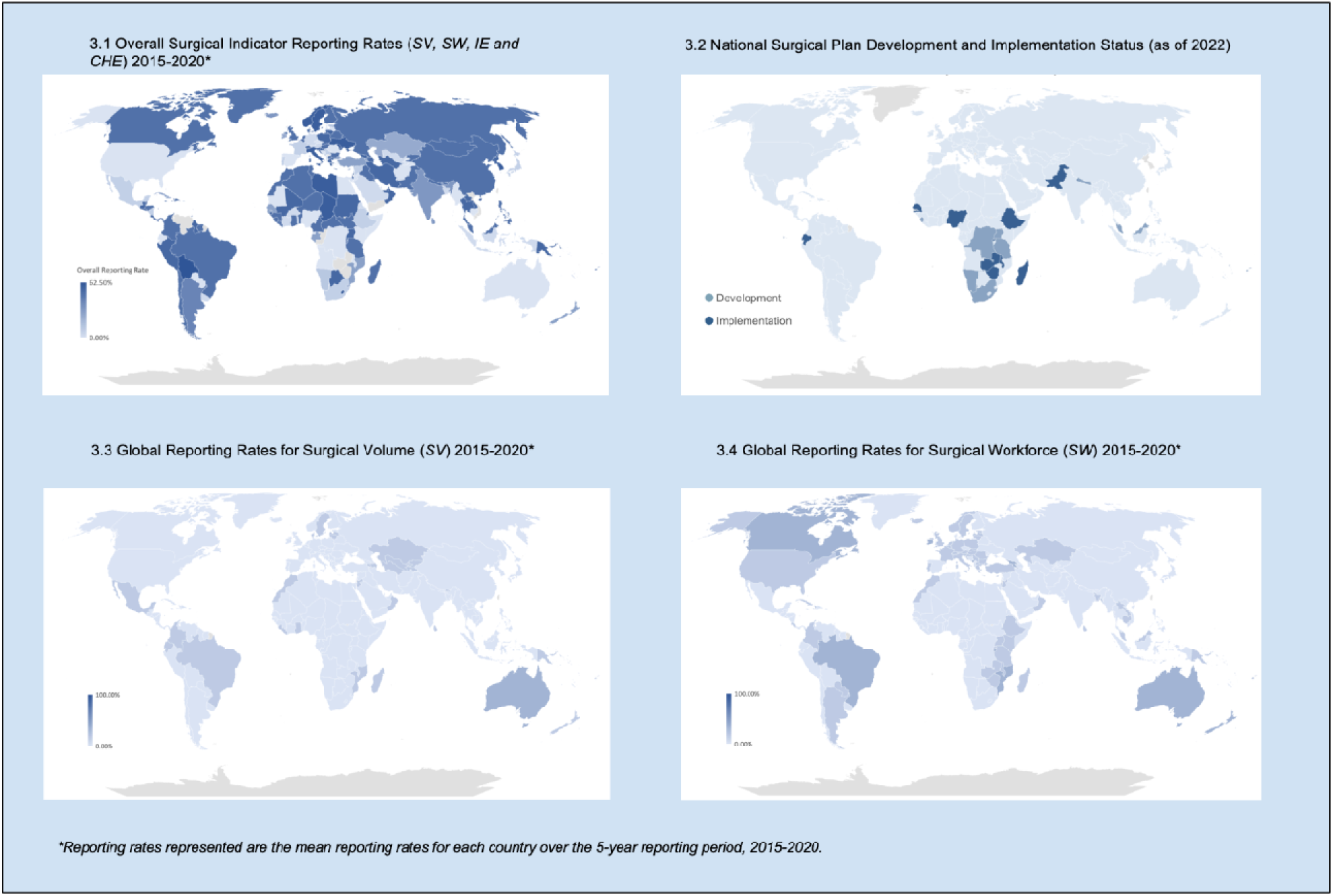
Global Trends in Surgical Indicator Reporting and National Surgical Plan Development and Implementation.

## DISCUSSION

The publication of the LCoGS catalyzed a global movement aimed at reducing disparities in access to safe, timely, affordable surgical care around the world. Since 2015, thirty-two countries have embarked on developing NSPs as frameworks to assess, prioritize, and strengthen their surgical health system, and eleven countries have successfully reached the implementation phase. (11) Despite the initial interest generated by the LCoGS, our analysis reveals notable gaps and a decline in the reporting rates of surgical indicators globally, with the causes behind this decline appearing to be multifactorial.

In the years immediately following the publication of the LCoGS, international collaboration research groups collected and compiled indicator data, subsequently providing it to the World Bank Group for publication. (12) Important considerations resulting from this initial work highlight some of the diverse challenges when attempting to collect indicator data. In addition to the well-described deficits in data availability and quality, financial indicators demonstrated additional obstacles secondary to the higher complexity in the collection of socioeconomic data, such as reliable data on household income and poverty thresholds, the estimation of direct and indirect costs (notably where costs and financial protections differ significantly between countries), and the lack of standard definitions for catastrophic and impoverishing expenditure. To address these challenges in primary data collection for the financial surgical indicators, Shrime et al. constructed a model to estimate these indicators. (13,14) The estimation of global data and reporting by a single group partially explains the higher reporting rates seen during the study period for the financial indicators. This demonstrates how continuous monitoring by a champion or group has the potential to improve indicator reporting (Box 2).

Additional barriers contributing to suboptimal indicator data collection include the lack of standardized reporting systems specific to surgical care data, deficient data infrastructure, and insufficient training in data management. (15–17) Attempts at collecting indicator data in the last decade have also uncovered challenges with health system governance, the variability in health information systems, contextual political factors, and the absence of participatory monitoring. (18,19). The resources needed for robust and accurate data collection and reporting systems appear disproportionately based in HICs, where both reporting of primary data and modeling occur at higher rates. Despite this, HICs, including the United States and Australia, have not consistently provided indicator data for indicator data for the WDI. The disparity between regional reporting rates underscores the need for targeted interventions to strengthen the capacity for surgical data collection in low-resource settings. No significant differences were noted in indicator reporting rates between countries that have embarked on developing a NSP and those that have not.

For the surgical volume indicator (SV), data are collected by the LCoGS and Center for Health Equity in Surgery and Anesthesia at UCSF Medical Center from various sources such as published academic journals, Ministry of Health, European Health For All Database, European Health Information Gateway, OECD database, and Eurostat. The collected data have been reported to the World Bank once every few years since 2016. If the data are extracted from published academic journals that are found through a structured search, the year of the source data may be some distant past. For the surgical workforce density indicator (SW), the data were collected by the LCoGS and WHO Collaborating Centre for Surgery and Public Health at Lund University from various sources including Ministries of Health, Medical Councils or Associations, Eurostat, health institutes, or government publications. The collected data were reported to the World Bank. However, this data collection system was discontinued after 2019 and currently there is no reporting system for the surgical workforce indicator. Therefore, the WDI has not had data updates for this indicator since then, and the WDI does not have any surgical workforce density data for 2019-2023. For financial risk indicators, updated modeled data are generated by the Program in Global Surgery and Social Change (PGSSC) at Harvard Medical School, and have been reported to the World Bank once every few years since 2016.

In some cases, nations will collect indicator data but may not reach the World Bank. Indicator data collected independently may have been published in academic journals or regional datasets but is absent in the WDI due to an unestablished reporting system. (20,21) By contrast, smaller-scale regional collaborations may more successfully incentivize data collection and sharing. The Western Pacific region (WPRO) offers a notable example, with health system leadership from multiple nations working together for strategic planning and knowledge sharing to strengthen surgical systems. (22)

Lessons can be drawn from the experiences of data collection for other indicators. For example, the International Diabetes Federation (IDF) provides the WDI data for global diabetes prevalence. This data is compiled using a combination of peer-reviewed publications and national health surveys. The IDF calculated estimates of diabetes prevalence for countries without available data using extrapolations from countries with similar characteristics. (24) For IDF metrics, data collection efforts face similar challenges of limited primary data availability encountered when compiling surgical indicators. Reliance on modeled data introduces numerous additional potential limitations and should not be used as a long-term substitute for primary data collection. (25) However, the consistency in reporting rates for the financial indicators CHE and IE demonstrates the role of extrapolation and modeling as a means of filling data gaps, especially while developing systems for primary data collection.

In 2021, the LCoGS surgical indicators were further refined with the Utstein consensus report, which provided more detailed descriptions of each indicator, defined the data points needed to calculate each indicator, and consolidated the two financial indicators into a single measure of catastrophic expenditure from surgical care. (7) These adjustments sought to make data collection and reporting more uniform and reliable, so that they could serve as a more realistic estimate of the reality of surgical access. Still, many low- and middle-income countries face significant resource constraints that impede effective data collection and reporting. (26)

Surgical systems are inherently complex, involving multiple stakeholders, diverse healthcare settings, and various levels of care. This complexity can make data collection and reporting challenging, particularly in limited-resource settings where healthcare infrastructure may be underdeveloped. The observed deficiencies in the reporting of surgical indicator data highlight the need for pragmatic, scalable, and sustainable mechanisms for collecting, reporting, and monitoring surgical care trends globally (Box 2). Improved data reporting is crucial for tracking progress and identifying gaps and areas needing intervention, ultimately leading to better health outcomes globally.

### Box 2

Recommendations for sustainable surgical indicator reporting

**Box 2: Recommendations for sustainable surgical indicator reporting**
| **Recommendation** | **Description** |
| --- | --- |
| ***Development of Standardized Data Collection and Reporting Mechanisms*** | Establish a standardized data collection framework for consistently collecting and reporting surgical indicators across all countries. These frameworks should be designed to accommodate the unique needs and capacities of different regions. Data, including metadata, should be reported to an international data collection system. The collected data should be reviewed and published. Through international collaboration, the sharing of best practices and resources for improving surgical care monitoring and reporting is made possible. This could involve partnerships between high-income and low-income countries, as well as collaboration with international organizations and non-governmental organizations. Another possible model includes using professional societies and advocacy groups to collect and collate surgical data from members and report them to WDI, lessening the burden on ministries. This is a model already in use by the World Bank for specific regions.<sup>21</sup> |
| ***Integration into NSPs*** | Adapt the existing NSP framework to include a clear strategic plan for surgical indicator collection. A pragmatic, funded framework for collection and reporting mechanisms enables countries to take ownership of their surgical data through the implementation of robust national reporting systems. This could involve strengthening leadership and governance at the national level to integrate surgical data reporting into the country's. Policymakers should drive data collection and reporting and invest in nationalized integrated data systems.(27) In addition to an integrated framework for indicator collection, a designated country lead for WDI surgical indicator reporting may facilitate better transparency and integration of surgical indicators into national priorities. |
| ***Sustainable Funding of Surgical Indicator Reporting*** | Invest in a strategic indicator collection plan and in capacity-building initiatives to improve countries' ability, particularly in regions with low reporting rates, to collect and report surgical data accurately. Surgical indicators can be used as outcome metrics to evaluate the impact of investments into surgical care delivery. Thus, investments in the reporting of surgical indicators have the potential to increase investment in the broader health system. Funding can be used to finance training programs for data collection and technical assistance or to develop health information systems. As innovations for global surgery financing take shape, data collection and indicator reporting strategies should be a core program for investment. |

### Limitations

The design of this study focuses on a descriptive analysis of overall patterns in reporting rates to the World Bank Group for presentation on its World Development Indicator platform. The methodology does not capture data collected and reported elsewhere, nor does it characterize collaborations between the WDI, data collectors, and national leadership. The relationship between data collection and reporting is complex and nuanced, and this manuscript reflects simply one facet of that reporting dynamic. This analysis reflects data availability as of May 2024 and therefore does not account for any data that may have been made available since then. With respect to national surgical plans, we acknowledge that countries may have national health policy addressing surgical care that was not retrieved with our search strategy and that the presence or absence of a plan does not speak specifically to the implementation phase, as well as the allocation of resources to data collection. While this project did not set out to describe the facilitators and barriers to indicator reporting, further research is needed in this area.

### Conclusion

Surgical indicator reporting rates to the World Development Indicators are low. Accurate and comprehensive data can inform policy decisions, guide resource allocation, and identify areas needing interventions. Further research is required to expand our collective understanding of specific barriers and potential optimizations to global surgical data collection.

## Data Availability

All data presented in the present study are available upon reasonable request to the authors. All primary data is freely available online.

https://databank.worldbank.org/source/world-development-indicators

## Author contributions

Conceptualization and design of the study: CT, KJ, VR, JGM, NPR

Literature review and synthesis: CT, KJ, VR

Initial data collection and management: CT, KJ

Methodology design: CT, KJ, VR, JGM, NPR

Quantitative statistical analysis: CT, KJ, VR

Final data cleaning, review and interpretation: CT, KJ, VR, GH, ES

Manuscript writing and revision: CT, KJ, VR, GH, ES

Review and editing of the manuscript: CT, KJ, VR, GH, JGM, NPR, ES

Final approval and revision: CT, KJ, VR, GH, JGM, NPR, ES

## Acknowledgments

nil

## Box 1

LCoGS indicators included in the World Development Indicator (WDI) dataset^1^

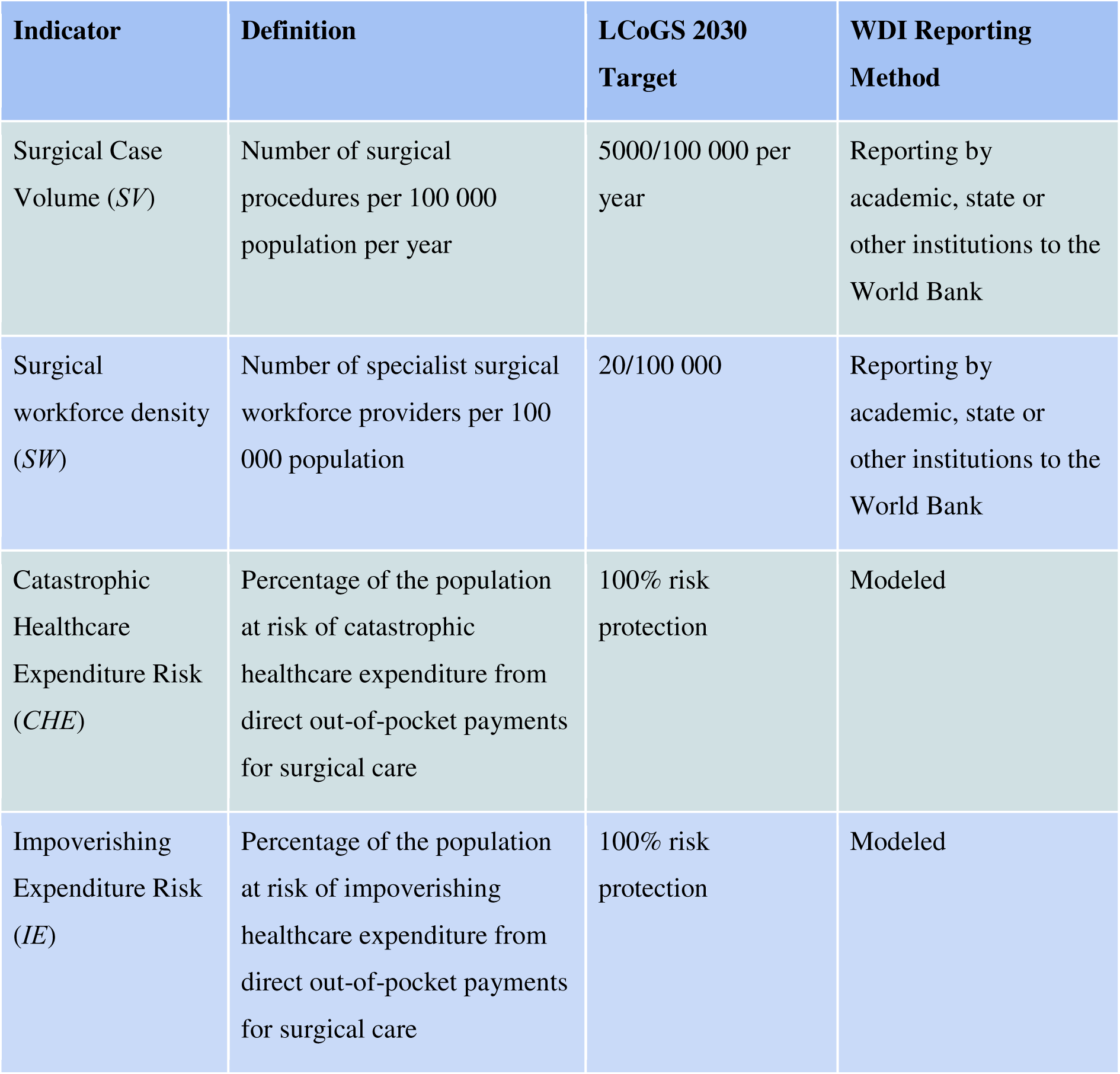

| Indicator | Definition | LCoGS 2030 Target | WDI Reporting Method |
| --- | --- | --- | --- |
| Surgical Case Volume (*SV*) | Number of surgical procedures per 100 000 population per year | 5000/100 000 per year | Reporting by academic, state or other institutions to the World Bank |
| Surgical workforce density (*SW*) | Number of specialist surgical workforce providers per 100 000 population | 20/100 000 | Reporting by academic, state or other institutions to the World Bank |
| Catastrophic Healthcare Expenditure Risk (*CHE*) | Percentage of the population at risk of catastrophic healthcare expenditure from direct out-of-pocket payments for surgical care | 100% risk protection | Modeled |
| Impoverishing Expenditure Risk (*IE*) | Percentage of the population at risk of impoverishing healthcare expenditure from direct out-of-pocket payments for surgical care | 100% risk protection | Modeled |

