## Supplemental Table 1 for "Tracking Progress: Trends in Surgical Indicator Reporting in the World Development Indicators"

**Supplementary material**

**Appendix 1: Regional surgical indicator data reporting rates from 2015 to 2020**

|  | **Number of surgical procedures (per 100,000 population)** | | | | | |
| --- | --- | --- | --- | --- | --- | --- |
|  | 2015 | 2016 | 2017 | 2018 | 2019 | 2020 |
| AFRO | 4.35% | 2.17% | 2.17% | 2.17% | 0.00% | 0.00% |
| EMRO | 14.29% | 0.00% | 0.00% | 0.00% | 0.00% | 0.00% |
| EURO | 20.37% | 0.00% | 1.85% | 0.00% | 0.00% | 0.00% |
| AMRO | 13.51% | 0.00% | 0.00% | 0.00% | 0.00% | 0.00% |
| SEARO | 18.18% | 9.09% | 0.00% | 0.00% | 0.00% | 0.00% |
| WPRO | 4.00% | 44.00% | 0.00% | 0.00% | 0.00% | 0.00% |
|  | **Risk of catastrophic expenditure for surgical care (% of people at risk)** | | | | | |
|  | 2015 | 2016 | 2017 | 2018 | 2019 | 2020 |
| AFRO | 78.26% | 76.09% | 76.09% | 78.26% | 78.26% | 76.09% |
| EMRO | 52.38% | 52.38% | 52.38% | 52.38% | 47.62% | 47.62% |
| EURO | 83.33% | 83.33% | 83.33% | 83.33% | 83.33% | 83.33% |
| AMRO | 62.16% | 62.16% | 62.16% | 62.16% | 62.16% | 62.16% |
| SEARO | 72.73% | 72.73% | 72.73% | 72.73% | 72.73% | 72.73% |
| WPRO | 40.00% | 40.00% | 40.00% | 40.00% | 40.00% | 40.00% |
|  | **Risk of impoverishing expenditure for surgical care (% of people at risk)** | | | | | |
|  | 2015 | 2016 | 2017 | 2018 | 2019 | 2020 |
| AFRO | 78.26% | 76.09% | 76.09% | 78.26% | 78.26% | 76.09% |
| EMRO | 52.38% | 52.38% | 52.38% | 52.38% | 47.62% | 47.62% |
| EURO | 83.33% | 83.33% | 83.33% | 83.33% | 83.33% | 83.33% |
| AMRO | 62.16% | 62.16% | 62.16% | 62.16% | 62.16% | 62.16% |
| SEARO | 72.73% | 72.73% | 72.73% | 72.73% | 72.73% | 72.73% |
| WPRO | 40.00% | 40.00% | 40.00% | 40.00% | 40.00% | 40.00% |
|  | **Specialist surgical workforce (per 100,000 population)** | | | | | |
|  | 2015 | 2016 | 2017 | 2018 | 2019 | 2020 |
| AFRO | 2.17% | 23.91% | 0.00% | 6.52% | 0.00% | 0.00% |
| EMRO | 9.52% | 4.76% | 4.76% | 0.00% | 0.00% | 0.00% |
| EURO | 44.44% | 9.26% | 14.81% | 11.11% | 0.00% | 0.00% |
| AMRO | 10.81% | 5.41% | 2.70% | 16.22% | 0.00% | 0.00% |
| SEARO | 9.09% | 9.09% | 0.00% | 9.09% | 0.00% | 0.00% |
| WPRO | 8.00% | 52.00% | 4.00% | 12.00% | 0.00% | 0.00% |
